# Electrical signatures of divergent connectivity in the human subgenual cingulate cortex

**DOI:** 10.64898/2026.06.09.26355288

**Authors:** Zekai Qiang, Panagiotis Kerezoudis, Nicholas Gregg, Dora Hermes, Bryan T Klassen, Aswin Chari, Martin M Tisdall, Matthew R Baker, Kai J Miller

## Abstract

**Background:** Major depressive disorder remains a leading cause of disability. While subgenual cingulate cortex (sgCC) deep brain stimulation (DBS) shows promise for medically refractory depression, clinical outcomes have been heterogeneous, suggesting that individual differences in neural circuitry engagement may critically influence therapeutic efficacy. We aimed to define the electrophysiological signatures of sgCC efferent connectivity using single-pulse electrical stimulation (SPES) with intracranial stereo-EEG (sEEG) to inform rational targeting and physiological biomarkers for sgCC-DBS.

**Methods:** In four patients undergoing clinically indicated sEEG for seizure mapping, SPES was delivered through sgCC pairs, while distributed brain stimulation-evoked potentials (BSEPs) were recorded across cortical and subcortical sites. Responses were characterized using Canonical Response Parameterization to extract reproducible waveforms and per-trial reliability.

**Results:** sgCC stimulation elicited reproducible, spatially organized BSEPs across frontal, limbic, and paralimbic networks, aligning with known anatomical pathways. Frontal recruitment featured robust, lateralized orbitofrontal activation favoring the ipsilateral central, medial OFC and bilateral ventromedial prefrontal responses. Limbic effects demonstrated bilateral cingulate activation with stronger ipsilateral recruitment and lateralized amygdala and hippocampal responses. Paralimbic engagement included insular responses with subject-specific anterior predominance and bi-hemispheric temporal-polar slow-wave deflections.

**Conclusion:** These findings provide direct electrophysiological evidence of distributed, lateralized sgCC divergent network connectivity in the human brain, offering physiologic confirmation of its role in affective circuitry. The observed topography and laterality have direct applications for sgCC-DBS targeting and implicate BSEP signatures as candidate biomarkers to guide patient-specific therapy.

## INTRODUCTION

Major depressive disorder (MDD) is among the leading causes of global disability, contributing substantially to morbidity and mortality. In the United States, nearly one in five adults report a lifetime diagnosis of depression (1). Despite the availability of effective pharmacologic and psychotherapeutic interventions, a large proportion of patients continue to experience persistent symptoms. Treatment-resistant depression (TRD), defined by inadequate response to at least two antidepressants of sufficient dosage and duration, affects an estimated 30-48% of individuals with major depressive disorder, representing a significant clinical challenge that demands innovative therapeutic approaches (2, 3).

Deep brain stimulation (DBS) has emerged as a promising intervention for TRD, delivering targeted high-frequency electrical stimulation to modulate pathological neural activity within distributed reward and affective circuits (4). Several candidate targets have been explored, including the subgenual cingulate cortex (sgCC), lateral habenula, ventral striatum, medial forebrain bundle, and inferior thalamic peduncle (5). Among these, the sgCC has been the most extensively studied, owing to its central role in integrating affective, cognitive, and autonomic processes relevant to depression. However, clinical outcomes following sgCC DBS have been heterogeneous, with reported response rates ranging from 23% to 92% (6, 7), suggesting that individual differences in neural circuitry engagement may be critical determinants of therapeutic efficacy.

An improved understanding of sgCC connectivity is therefore essential for refining DBS targeting and stimulation paradigms. Structural tractography and diffusion imaging have delineated sgCC connections to key limbic and frontal regions, including the nucleus accumbens, amygdala, hypothalamus, and orbitofrontal cortex (8). Recent connectomic-guided DBS approaches targeting the intersection of the cingulum bundle, forceps minor, uncinate fasciculus, and frontostriatal fibers have yielded markedly improved outcomes up to 81.8% at one year (9, 10), emphasizing the importance of precise white matter engagement. Yet, these methods remain inferential, providing correlational rather than causal evidence of human sgCC network connectivity.

Direct electrophysiological mapping of sgCC connectivity has been limited. Prior work using scalp electroencephalography has identified characteristic stimulation-locked cortical responses following chronic sgCC DBS, implicating orbitofrontal and posterior cortical regions (11, 12), while local field potential studies reveal dynamic beta-band modulation during treatment (13). However, these approaches lack the spatiotemporal precision to resolve directional, region-specific propagation of activity within deep and cortical structures.

Single-pulse electrical stimulation (SPES) delivered through intracranial stereo-electroencephalography (sEEG) provides a unique opportunity to directly probe causal connectivity with millisecond precision. In this study, we applied SPES to the sgCC in patients undergoing sEEG implantation for seizure localization. By recording brain stimulation-evoked potentials (BSEPs) across distributed cortical and subcortical regions, we sought to delineate the electrophysiological signatures of sgCC connectivity in the human brain(14, 15). These findings provide causal evidence for the distributed circuits engaged by sgCC stimulation, informing future strategies for rational DBS targeting and optimization in depression.

## METHODS

### Ethics Statement

This study was approved by the Mayo Clinic Institutional Review Board (IRB #15-006530) and conducted in accordance with the Declaration of Helsinki. All participants were undergoing intracranial monitoring for clinical purposes, and no modifications to clinical procedures were made for research. Each participant or their legally authorized representative provided written informed consent to participate in the study and for the use and sharing of their de-identified data as approved by the IRB.

### Subjects

Four patients undergoing sEEG implantation for seizure network characterization in the evaluation of drug-resistant epilepsy participated in this study. Electrode implantation sites and trajectories were determined solely on clinical grounds by the treating epilepsy team, and no additional electrodes were placed for research purposes(16, 17). All structural MRI images were defaced prior to analysis and data sharing to ensure participant anonymity.

### Lead Placement and Localization

sEEG leads were implanted according to standard clinical protocols for seizure network mapping. Depth electrodes (DIXI Medical) consisted of 10-18 platinum-iridium contacts, each 2mm in length and 0.8mm in diameter, with 1.5mm spacing between contacts (**Figure. 1A**). Electrode localization was performed by co-registering the post-implantation CT scan to the pre-implantation T1-weighted MRI. Each preoperative MRI was aligned to the anterior– posterior commissure (AC–PC) coordinate space, and the post-implant CT was co-registered to this MRI using mutual information implemented in SPM12 (18).

**Figure 1.**
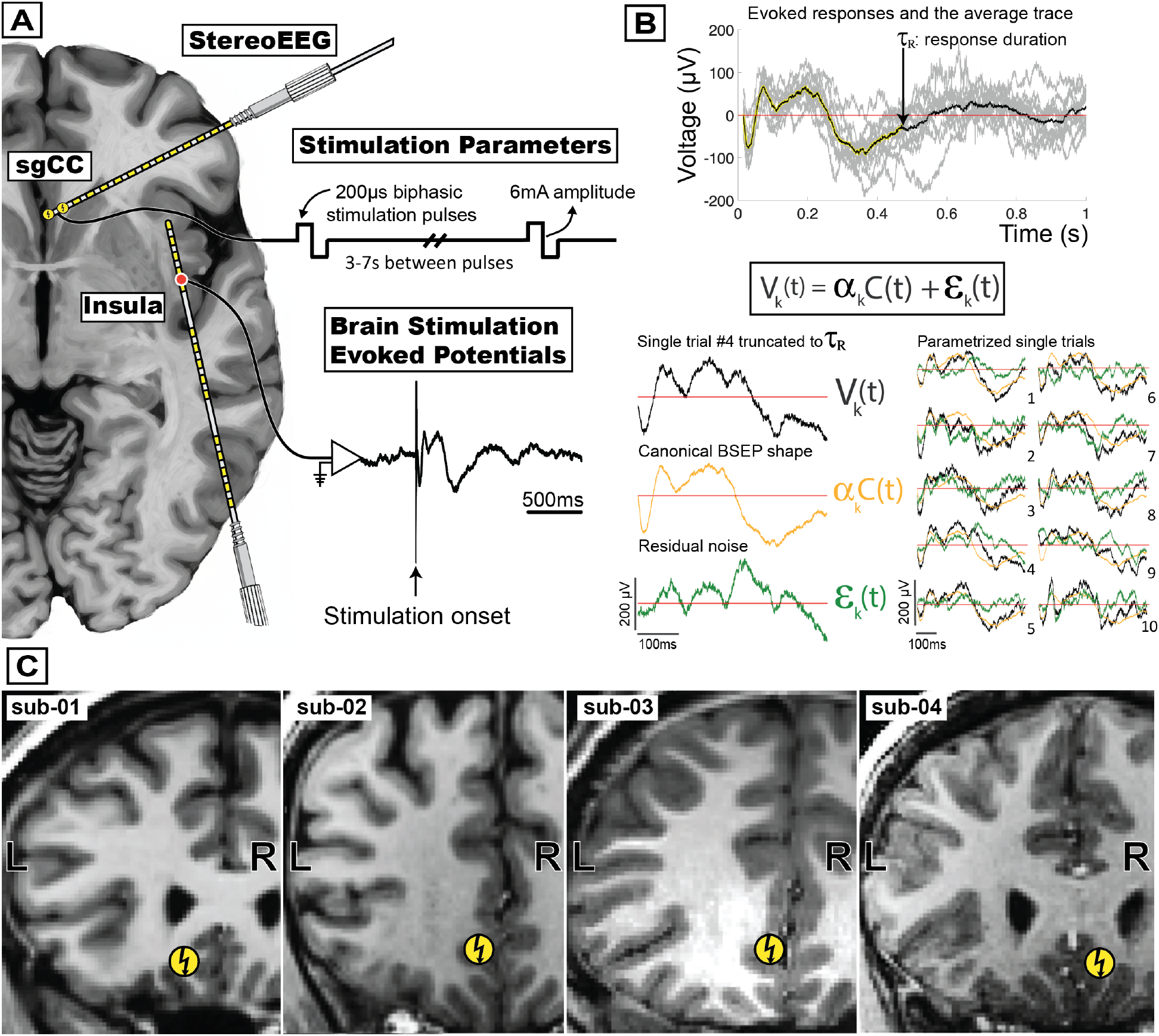
Single pulse electrical stimulation and canonical response parameterization. **A**. A subgenual cingulate electrode pair delivered 12 single biphasic pulses (6 mA, 100 µs) with randomized interstimulus intervals of 3–7 s. **B**. The canonical response parameterization method models each stimulation-evoked trial as *V*_*k*_(*t*) = *α*_*k*_ *C*(*t*) + *ε*_k_(*t*), where *C*(*t*) is a canonical response waveform, *α*_*k*_ is a scalar amplitude, and *ε*_k_(*t*) is residual noise. **C**. Stimulation sites for each participant (n = 4) in the current study are shown.

### Single Pulse Electrical Stimulation

All study procedures were conducted in the Epilepsy Monitoring Unit or the Pediatric Intensive Care Unit at Mayo Clinic, Rochester, Minnesota. Stimulation sites were chosen for each participant according to prespecified clinical hypotheses. Each selected electrode pair delivered 10 to 12 stimuli consisting of single 6 mA, 100 µs biphasic pulses with randomized 3–7 s interstimulus intervals, administered using a g.Estim Pro stimulator (g.Tec, Schiedlberg, Austria). Voltage recordings were acquired with a g.HighAmp amplifier at 2400 Hz in three patients and 4800 Hz in one patient, with a white matter contact serving as the hardware ground. Trials contaminated by artifacts or epileptiform activity were identified and excluded prior to analysis. An epileptologist continuously supervised all stimulation sessions to detect ictal activity, and seizure rescue medications were available should stimulation precipitate a seizure. Subjects were included if at least one stimulation site was localized to the subgenual cingulate gyrus, that is, immediately inferior to the genu and caudal rostrum of the corpus callosum. **(Figure 1C)**

### Signal Processing

The recorded BSEPs were preprocessed with a high-pass filter configured with a 0.5 Hz passband, 0.05 Hz stopband, and 3 dB passband ripple, followed by notch filtering at 60 Hz and its first two harmonics using a fourth-order, zero-phase Butterworth design. After filtering, data were re-referenced using the bipolar montage by computing the voltage difference between immediately adjacent contacts, yielding differential channels that emphasize shared local activity between paired recording sites while suppressing common-mode noise; only directly neighboring pairs were formed.

### Canonical Response Parameterization

The Canonical Response Parameterization (CRP) method was applied to BSEPs from 15 ms to 1 s after stimulation to minimize contamination from direct stimulation artifacts and volume conduction. For each stimulation–recording site pair, this data-driven approach identifies reproducible response morphologies and, when present, estimates response duration based on a temporal profile of structural significance, rather than imposing a predefined waveform shape. (**Figure 1B**) We refer the readers to the original paper for full methodological details (15). Individual trial parameters are then extracted, including projection coefficient *α*_*k*_ = ∑_*t*_ *C*(*t*) *V*_*k*_(*t*) amplitude normalized by 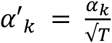, explained variance 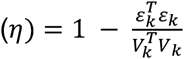. The coefficient of determination (*R*^2^) was computed on per-trial basis as the mean subtracted explained variance according to 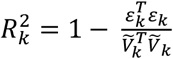. Values of *R*^2^ near 0 indicate that *C*(*t*) explains little of the inter-trial variability, whereas values near 1 indicate that *C*(*t*) explains almost all the variability and thus there is highly reliable connectivity. Statistical significance for each stimulation–recording interaction was assessed using extraction significance based on one-tailed t-test with an alpha threshold of 0.05.

### Electrode Visualization and Anatomical Labeling

For visualization, channels were displayed on axial, coronal, and sagittal T1-weighted MRI planes using the SEEGVIEW package, which samples slices at fixed intervals and projects each contact to the nearest selected slice to present responses and anatomical locations in a clinically interpretable format (19). Because projection distance increases with slice thickness, a true gray matter site may artifactually appear in adjacent structure after projection, and vice versa. Marker size and color intensity for each channel were jointly scaled to the maximum weight of 1, and channels deemed insignificant (that is, extraction significance >0.05) were rendered as small, gray circles of fixed diameter. For each subject, T1 MRI scans were segmented with the built-in auto-segmentation pipeline in FreeSurfer 7 (20). Channel-level anatomical labels were then derived from subject-specific gyral and sulcal anatomy using the Destrieux cortical atlas (21).

## RESULTS

Across four subjects, SPES of the sgCC elicited reproducible and spatially organized BSEPs spanning frontal, limbic, and paralimbic networks (**Figure 2, Suppl Table 1**). Response consistency, quantified by the coefficient of determination (*R*^2^), varied across regions and hemispheres, but revealed activation within circuits known to mediate affective and autonomic regulation (22).

**Figure 2.**
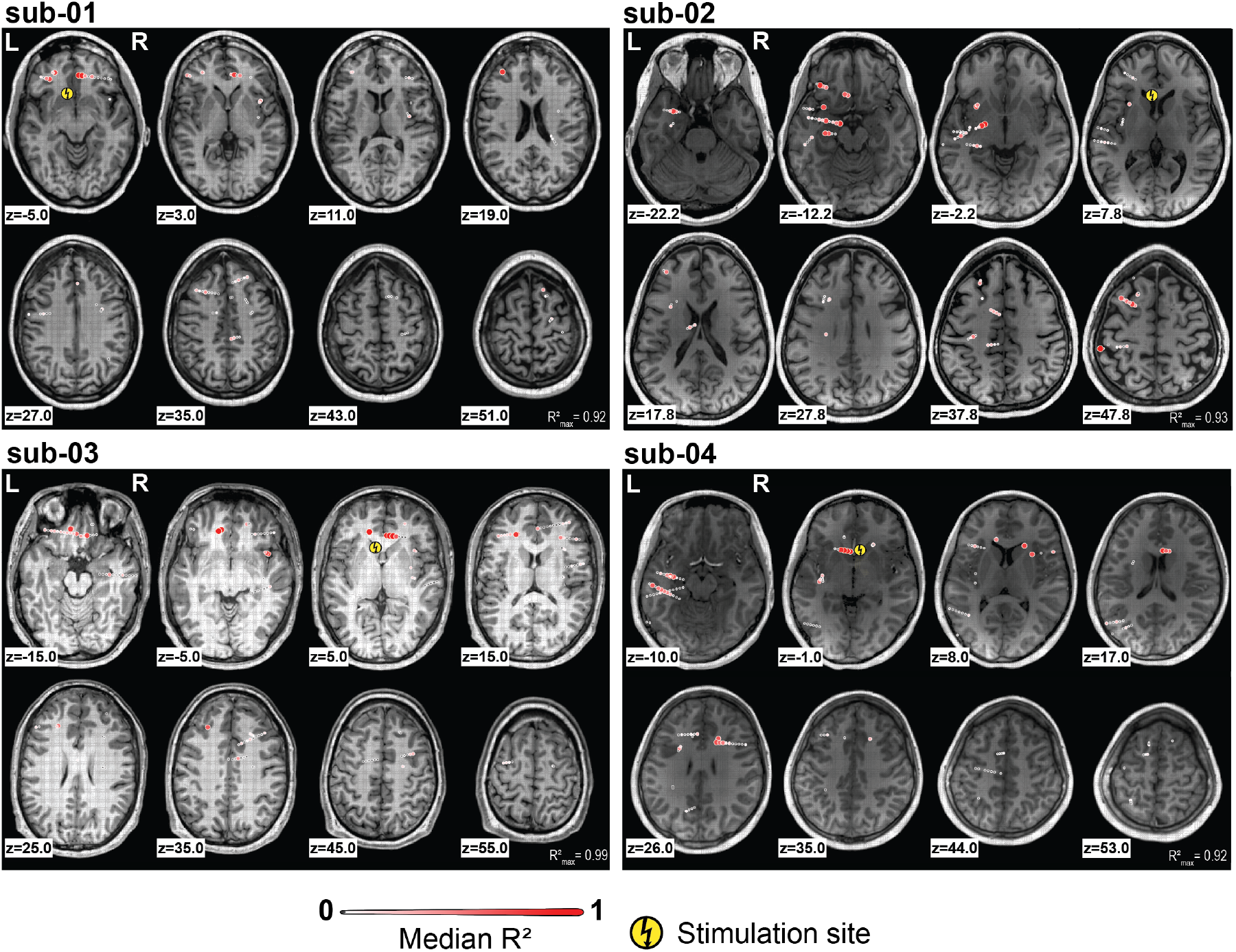
Global activation patterns after single pulse electrical stimulation of the subgenual cingulate cortex. Axial T1-weighted slices are shown for four individual subjects 1 to 4. Distinct activation patterns are observed in frontal, limbic, and paralimbic networks. Greater values of *R*^2^ indicate more reliable causal connectivity estimates, with 0 denoting no reliable relationship and 1 representing maximally reliable connectivity.

### Frontal Network

Bilateral orbitofrontal cortical recordings were available in two participants (subjects 1 and 3). In both cases, ipsilateral sgCC stimulation elicited more robust orbitofrontal responses than contralateral stimulation (Subject 1: ipsilateral number of significant channels 4/5 (80%), significant channels median R^2^ 0.57; contralateral number of significant channels 2/9 (22.22%), significant channels median R^2^ 0.42; p = 0.038. Subject 2: ipsilateral number of significant channels 8/10 (80%), significant channels median *R*^2^ 0.22, contralateral number of significant channels 4/17 (23.53%), significant channels median R^2^ 0.18; p = 0.005), suggesting a lateralized engagement of orbitofrontal circuitry. The most robust effects were localized to the central and medial orbitofrontal cortex (**Figure 3A, Suppl Figure 1)**.

**Figure 3.**
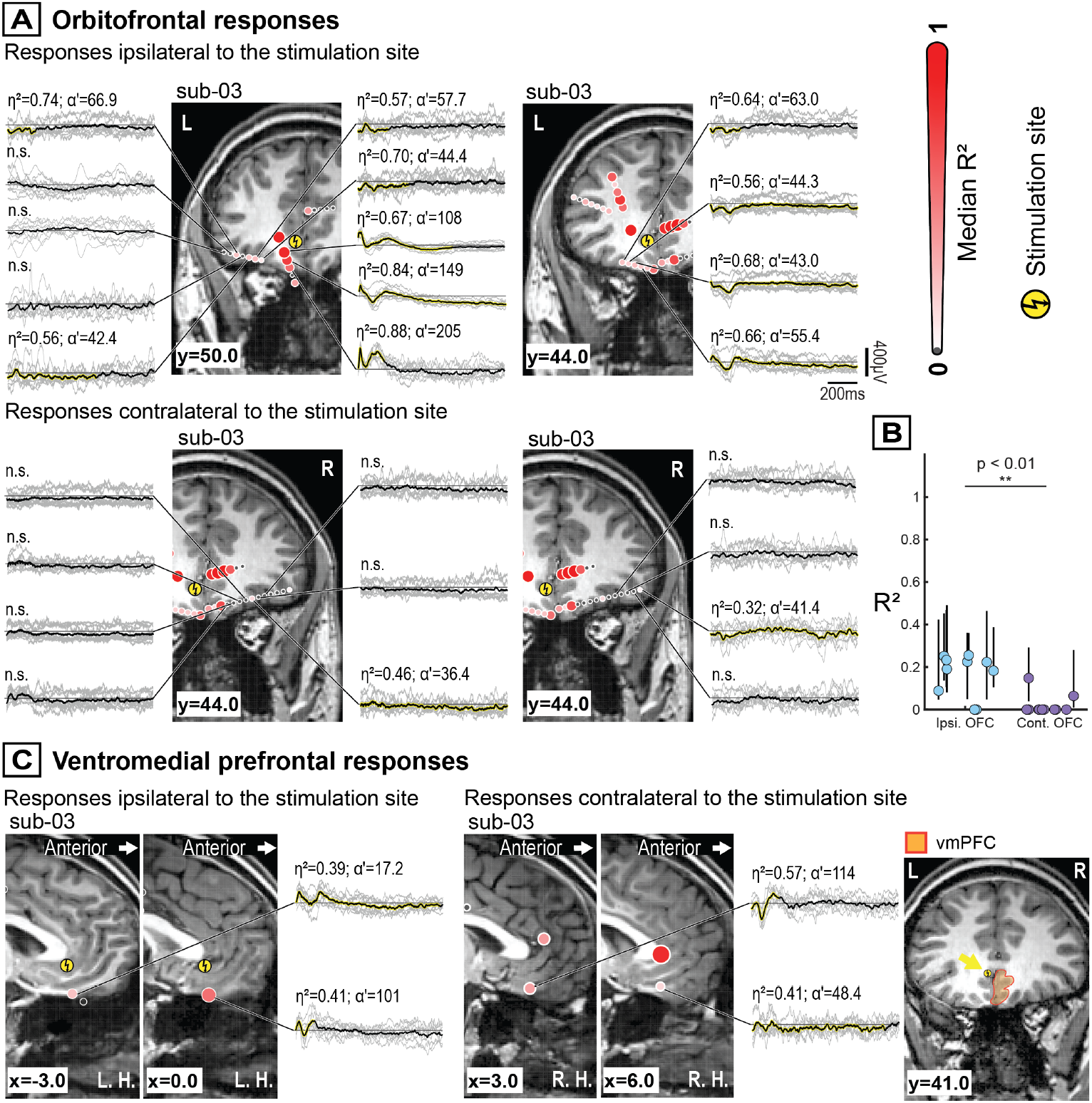
Frontal network responses following subgenual cingulate stimulation. **A**. Ipsilateral stimulation produced stronger engagement of the orbitofrontal (OFC) circuit compared with contralateral stimulation. **B**. In subject 3, significantly greater (*R*^2^) values were observed in the OFC ipsilateral to the stimulation site. Each dot represents the median (*R*^2^) across trials for a recording channel, with error bars indicating the interquartile range (Mann–Whitney U test, p<0.01). **C**. Unilateral subgenual cingulate stimulation elicited bilateral activation within the ventromedial prefrontal cortex in subject 3.

Ventromedial prefrontal cortical (vmPFC) recordings were available in three subjects and exhibited bilateral activation following sgCC stimulation. In subject 3, there was no significant difference in *R*^2^ between contralateral and ipsilateral vmPFC evoked responses (ipsilateral number of significant channels 4/4 (100.00%), significant channels median R^2^ 0.66; contralateral number of significant channels 3/4 (75.00%), significant channels median R^2^ 0.36; p = 0.038) (**Figure 3B, Suppl Figure 2**).

### Limbic Network

The cingulate cortex can be partitioned into anterior and posterior divisions, where the anterior cingulate is defined as the portion situated rostral to the vertical anterior commissural line, whereas the posterior cingulate occupies territory caudal to this landmark. Unilateral stimulation of the sgCC elicited bilateral responses in the cingulate cortex. Anterior and posterior responses in the cingulate cortex upon contralateral stimulations were available in three subjects, and they did not exhibit a consistent response topography (Subject 1 contralateral stimulation: anterior number of significant channels: 4/5 (80.00%), significant channels median *R*^2^ 0.23; posterior number of significant channels: 2/7 (28.57%), significant channels median *R*^2^ 0.30, p = 0.250). (**Figure 4A, Suppl Figure 3**)

**Figure 4.**
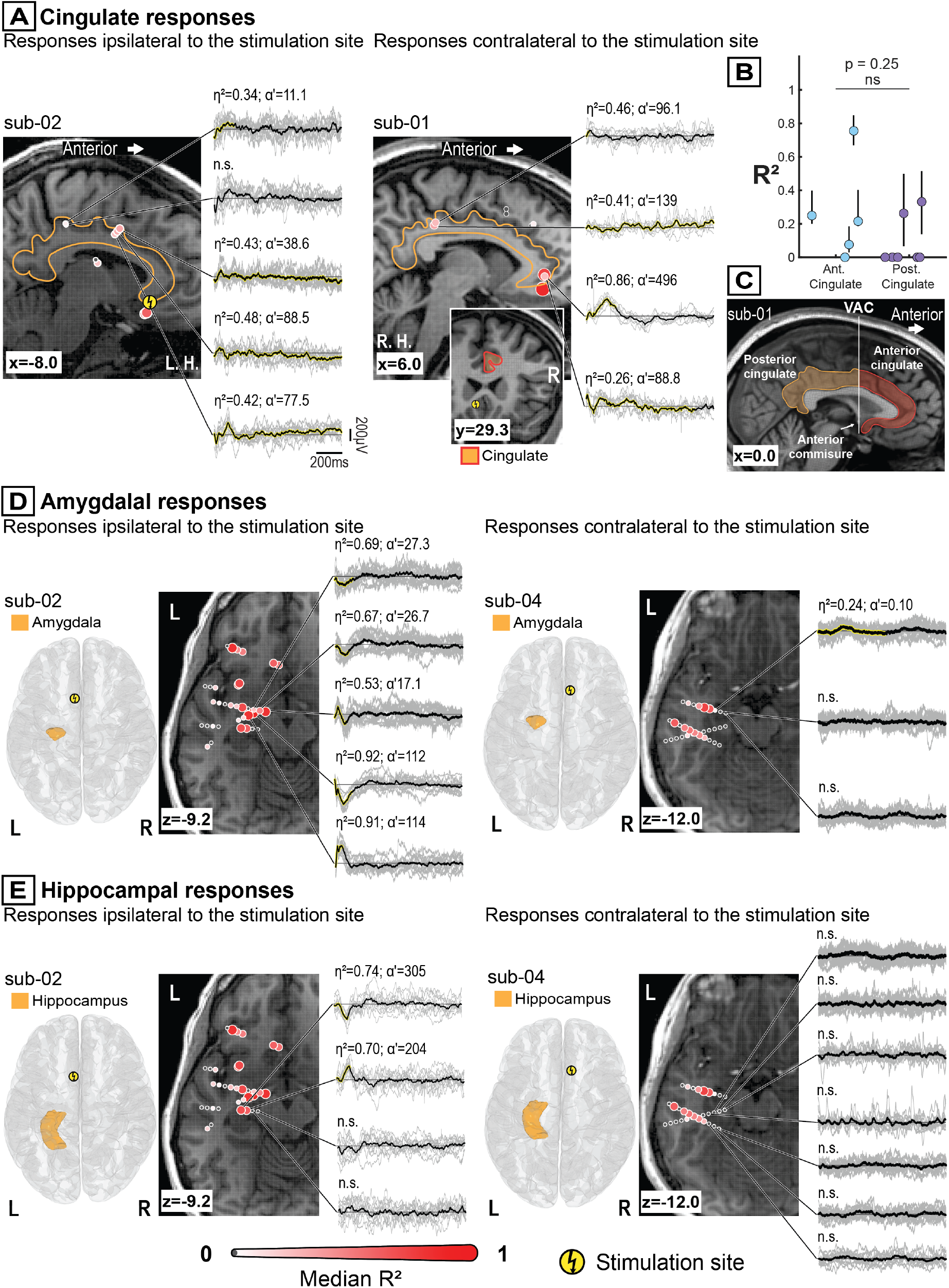
Limbic network responses following subgenual cingulate stimulation. **A**. Unilateral subgenual cingulate (sgCC) stimulation evoked bilateral cingulate activity. **B**. No significant difference in connectivity was observed between the anterior and posterior cingulate following stimulation of the contralateral sgCC. Each dot represents the median value across trials for an individual recording channel, with error bars indicating the interquartile range (Mann–Whitney U test, p=0.25). **C**. The vertical anterior commissural (VAC) line is interpolated perpendicular to the anaterior commissure and divides the cingulate cortex in anterior and posterior portions. **D**. The ipsilateral amygdala showed robust responses to stimulation, while contralateral amygdala sites exhibited minimal activity. **E**. The hippocampus demonstrated similarly lateralized engagement, with stronger ipsilateral than contralateral responses to sgCC stimulation.

In a single subject 2, the ipsilateral amygdala showed a stronger response than the contralateral side, suggesting possible lateralized engagement in this individual. In subject 3 and 4, contralateral amygdala sites are largely weak or non-significant (**Figure 4B**). Similarly, the hippocampus exhibits stronger ipsilateral than contralateral responses after sgCC stimulation. (**Figure 4C, Suppl Figure 4**)

### Paralimbic Network

sgCC stimulation reliably evoked BSEPs within the insula in both ipsilateral and contralateral regions. The insula is divided by the central insular sulcus into anterior and posterior portions (23). In the single subject 2 where ipsilateral recordings were available, sgCC stimulation produced stronger responses in the anterior than the posterior insula (Subject 2: anterior number of significant channels: 6/7 (85.71%), significant channels median *R*^2^ 0.42; posterior number of significant channels: 4/9 (44.44%), significant channels median *R*^2^ 0.14, p = 0.023).

Contralateral insular recordings were available in three participants; among these, subject 3 showed stronger anterior contralateral insular activation when the sgCC gray matter was stimulated (Subject 3: anterior number of significant channels: 7/7 (100.00%), significant channels median *R*^2^ 0.25; posterior number of significant channels: 3/7 (42.86%), significant channels median *R*^2^ 0.09, p = 0.009). However, this effect not observed in subject 1 (white-matter stimulation, anterior number of significant channels: 4/8 (50.00%), significant channels median *R*^2^ 0.17; posterior number of significant channels: 1/2 (50.00%), significant channels median *R*^2^ 0.30, p = 0.889) or subject 4 (gray–white junction, anterior number of significant channels: 1/7 (14.29%), significant channels median *R*^2^ 0.33; posterior number of significant channels: 4/9 (44.44%), significant channels median *R*^2^ 0.24, p = 0.381). (**Figure 5A, Suppl Figure 5**) Temporal-polar engagement was observed in both ipsilateral and contralateral hemispheres, characterized by low-frequency deflections. (**Figure 5B**)

**Figure 5.**
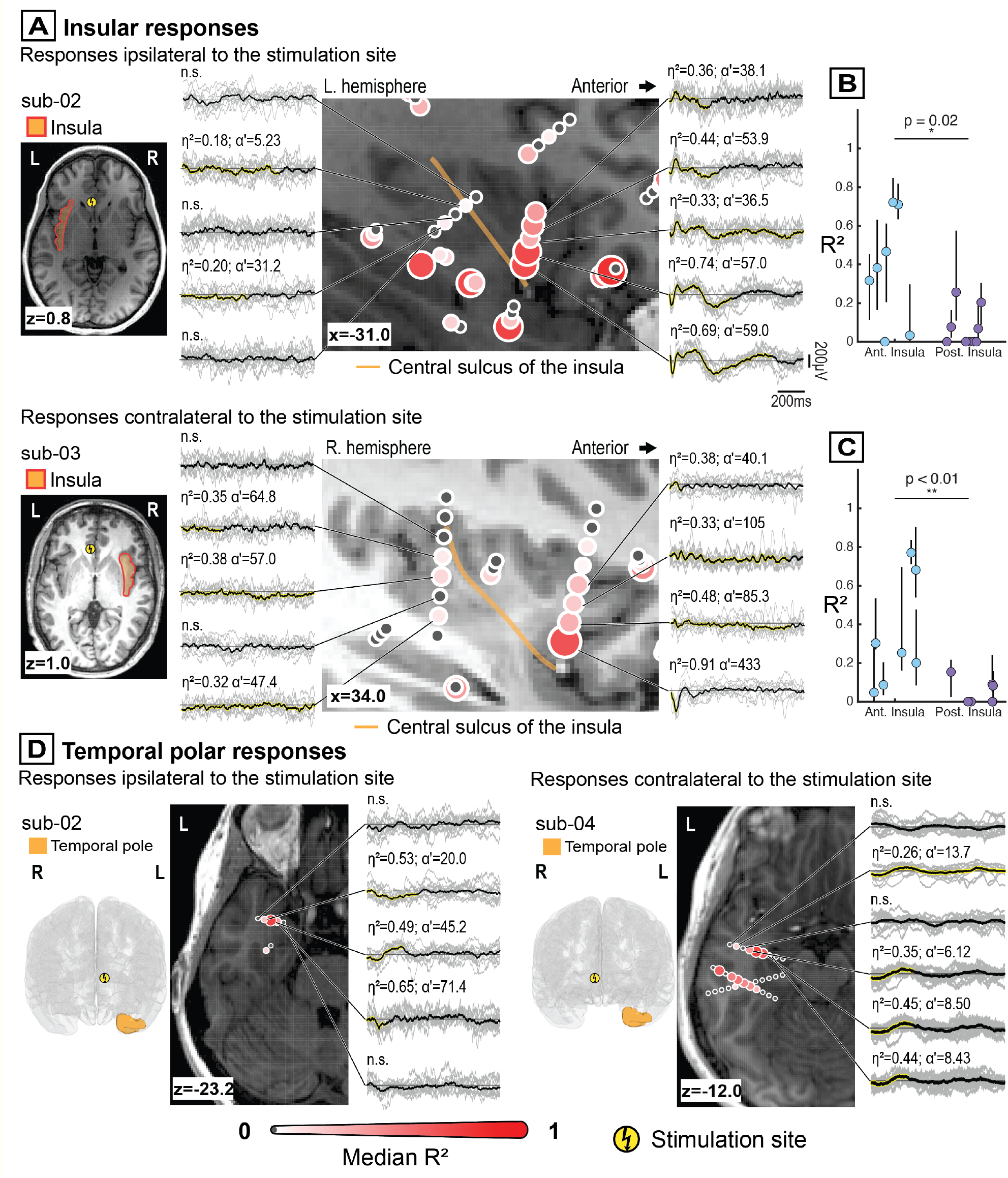
Paralimbic network responses following subgenual cingulate stimulation. **A**. subgenual cingulate (sgCC) stimulation elicited brain stimulation evoked potentials (BSEPs) within both ipsilateral and contralateral insula, with anterior insular responses appearing more prominent than posterior. **B**. The anterior insula exhibited significantly more reliable connectivity with the ipsilateral sgCC than the posterior insula. Each dot represents the median (*R*^2^) across trials for an individual recording channel, with error bars indicating the interquartile range (Mann–Whitney U test, p=0.02). **C**. In subject 3, the anterior insula exhibited significantly more reliable connectivity with the contralateral sgCC than the posterior insula. Each dot represents the median (*R*^2^) across trials for a recording channel, with error bars indicating the interquartile range (Mann–Whitney U test, (p=0.02)). **D**. Temporal pole engagement was observed bilaterally and was characterized by low-frequency, slow-wave activity.

## DISCUSSION

Single-pulse stimulation of the sgCC elicited reproducible, spatially organized BSEPs across frontal, limbic, and paralimbic territories, aligning with known anatomical pathways and delineating a functional architecture subserving affective regulation. These responses provide direct electrophysiological evidence of sgCC connectivity in humans, complementing inference from animal tract-tracing and diffusion models that left its causal organization unresolved. Together, these measurements position the sgCC as a central hub within distributed mood-regulatory networks and offer mechanistic context for its therapeutic relevance in depression neuromodulation.

Frontally, sgCC stimulation prominently recruited the OFC and vmPFC, regions known to have complementary roles in affective valuation and top-down control (24, 25). OFC responses were robust, spatially clustered, and lateralized toward the ipsilateral hemisphere, with preferential engagement of central and medial territories, demonstrating an organization consistent with projections via the cingulum and forceps minor that link the sgCC to these OFC subregions (8, 24). The OFC integrates multimodal sensory information and encodes the reward value of stimuli, providing a substrate through which sgCC output may influence behavioral motivation and hedonic appraisal (26). In contrast, the vmPFC which is involved in emotional regulation with extensive reciprocal connectivity involving the amygdala, anterior cingulate, and hypothalamus, exhibited bilateral activation (27). Together, the pattern of OFC and vmPFC activation delineates a ventral prefrontal pathway through which sgCC may recalibrate the balance between emotional salience and regulatory control, potentially explaining the antidepressant efficacy of sgCC-targeted stimulation.

Within limbic circuits, sgCC stimulation evoked organized responses in the cingulate, hippocampus, and amygdala, highlighting deep integration across emotion and memory systems. The anterior cingulate cortex demonstrates strong engagement consistent with its roles in affective evaluation, autonomic regulation, and conflict monitoring (28, 29), whereas more posterior cingulate territories, connected with retrosplenial and parietal regions, showed weaker or more variable effects (30). In addition, bilateral cingulate activation in response to unilateral sgCC stimulation may indicate that the sgCC has cingulate projections to both hemispheres or that homologous cingulate regions are connected via transcallosal pathways.

In the medial temporal lobe, amygdala and hippocampal responses were predominantly ipsilateral, mirroring primate tract-tracing that shows dense sgCC afferents, especially to medial and basolateral amygdala subregions (31). Given the amygdala’s role in emotional salience and the hippocampus’s role in contextual memory, these pathways may position the sgCC to couple affective appraisal with memory and autonomic output. Notably, reduced sgCC–amygdala coupling has been linked to impaired emotion regulation and depression severity in functional imaging, suggesting these pathways as plausible mediators of sgCC DBS effects (32-34).

Paralimbic engagement encompassed the insula and temporal pole, integrating interoceptive, emotional, and contextual representations. The insula exhibited subject-specific but potentially topographically organized responses, with anterior subdivisions showing stronger activation than posterior regions. This topography aligns with structural connectivity between the sgCC and anterior insula via the uncinate fasciculus and with salience network engagement that prioritizes evaluation of emotionally relevant internal and external signals (35-37). In contrast, posterior insular regions, which are more tightly coupled to somatosensory cortices, showed weaker or less consistent responses, suggesting preferential sgCC influence on affective rather than sensory facets of interoception (38). The temporal pole exhibited bi-hemispheric, low-frequency slow-wave deflections, consistent with its integrative role linking semantic and autobiographical memory to emotional and social meaning (39, 40).

## Conclusion

sgCC stimulation engaged a distributed network spanning frontal, limbic, and paralimbic regions that collectively support affective valuation, interoceptive awareness, and emotional memory. This systems-level organization provides a mechanistic framework for sgCC neuromodulation in depression and a roadmap for targeting network elements for therapeutic response. Methodologically, SPES maps causal human brain connectivity with high temporal precision, and the reproducibility of evoked responses across participants supports their robustness. At the same time, translating SPES-evoked dynamics into DBS optimization will require establishing clear links between these BSEP features and clinically meaningful outcomes, and ultimately demonstrating that modulating these responses alters depressive phenotypes. In this sense, SPES can be viewed as a candidate tool for patient-specific circuit characterization that may inform hypothesis-driven targeting strategies, pending prospective clinical validation. Key limitations of the present study include a small cohort (n = 4) of patients with clinically determined electrode placements, leading to heterogeneous sampling density, electrode orientation, gray–white matter engagement, and consequently variable circuitry coverage. Future work should integrate SPES-derived connectivity with multimodal structural and functional imaging to construct comprehensive causal connectomes of the sgCC, extend sampling across broader cortical territories in larger multicenter cohorts, and prospectively relate acute stimulation-evoked dynamics to chronic DBS outcomes.

## Data Availability

All data and code associated with the present study will be made available online with the final version of the article.

## Acknowledgments

This work was supported by the National Institutes of Health (NIH) NINDS U01-NS128612 (MRB, KJM) and R01-MH122258 (to D.H). Manuscript contents are solely the responsibility of the authors and do not necessarily represent the official views of the NIH. MRB and KJM are also supported by the Tianqiao & Chrissy Chen Institute. The funders had no role in study design, data collection and analysis, decision to publish, or preparation of the manuscript. ChatGPT (OpenAI) was used for assistance in text editing but not content generation.

## Supplementary Figures

**Supplementary Figure 1.**
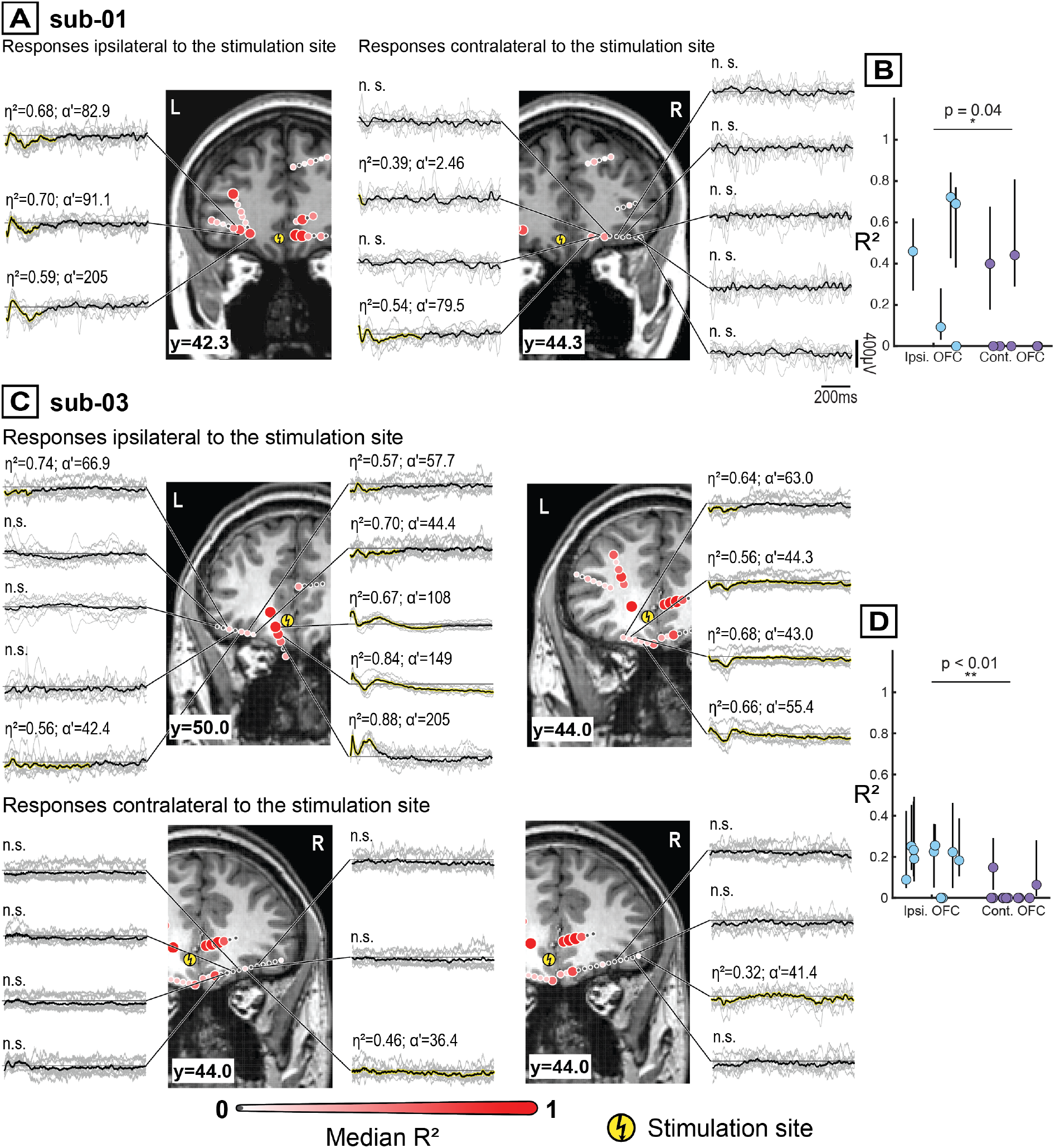
Orbitofrontal responses after subgenual cingulate stimulation. **A**. Brain stimulation evoked potentials (BSEPs) recorded from ipsilateral and contralateral orbitofrontal (OFC) electrodes following subgenual cingulate (sgCC) stimulation in subject 1. Significant responses were predominantly observed ipsilateral to the stimulation site, with non-significant responses contralaterally. **B**. Ipsilateral OFC contacts showed significantly higher *R*^2^ values than contralateral contacts (Mann–Whitney U test, p=0.04). Individual data points represent the median *R*^2^ across all trials in a recording channel with error bars indicating interquartile range. **C**. BSEPs recorded from ipsilateral and contralateral OFC contacts following sgCC stimulation for subject 3. Multiple ipsilateral OFC contacts showed significant responses, wheras contralateral OFC contacts showed predominantly non-significant responses and exhibiting low-amplitude waveforms. **D**. In subject 3, Ipsilateral OFC contacts showed significantly higher *R*^2^ values compared to contralateral OFC contacts (Mann–Whitney U test, p<0.01), consistent with an asymmetric, predominantly ipsilateral propagation of evoked activity from the sgCC to the OFC.

**Supplementary Figure 2.**
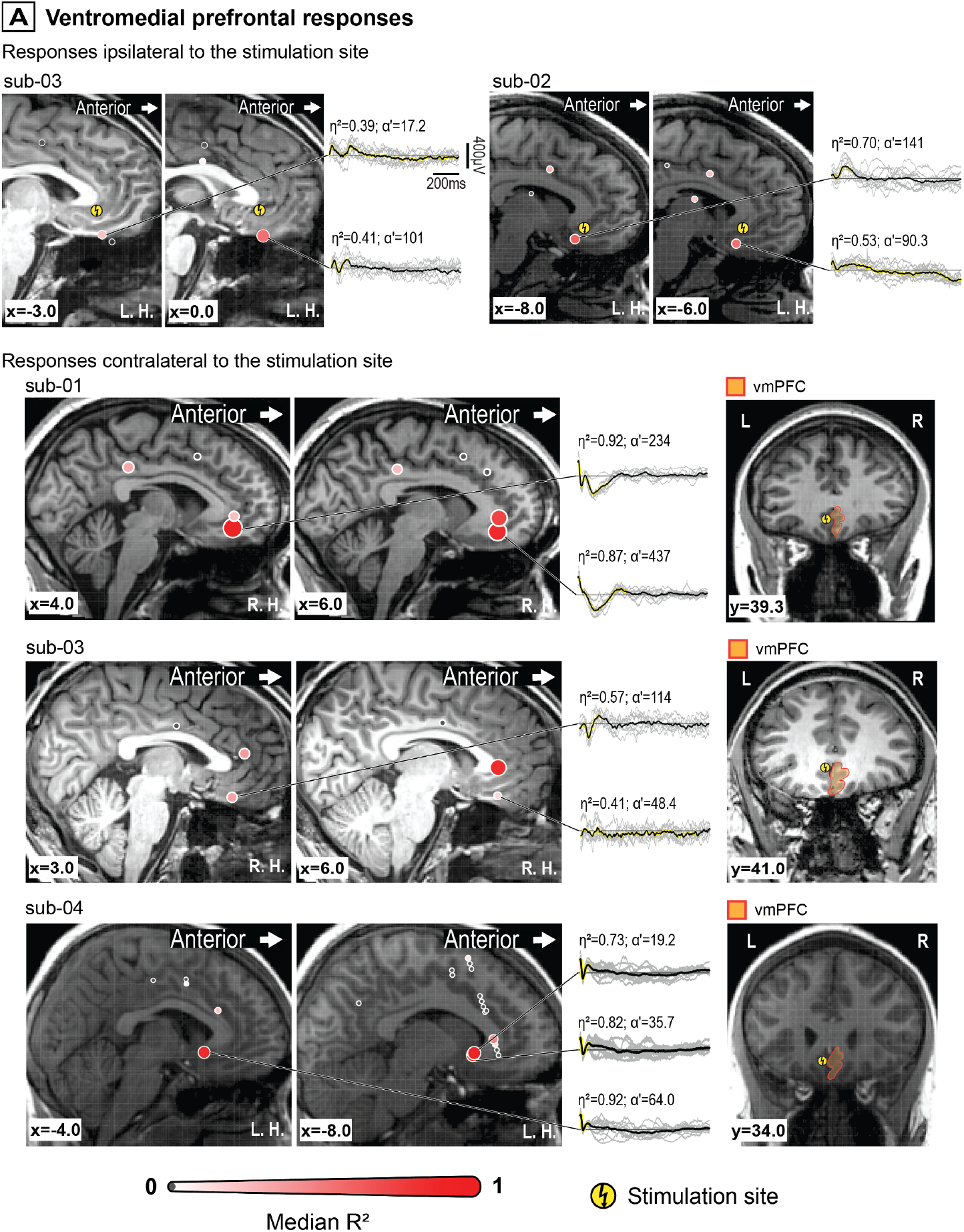
Ventromedial prefrontal responses after subgenual cingulate stimulation. **A**. Brain stimulation evoked potentials (BSEPs) recorded from ventromedial prefrontal cortex (vmPFC) electrodes ipsilateral to the subgenual cingulate cortex (sgCC) stimulation site in subjects 2 and 3. Multiple ipsilateral vmPFC contacts exhibited significant responses. For BSEPs recorded from vmPFC electrodes contralateral to the sgCC stimulation site in subjects 1, 3, and 4, significant responses were also observed in several contralateral vmPFC contacts, demonstrating bilateral propagation of stimulation-evoked activity.

**Supplementary Figure 3.**
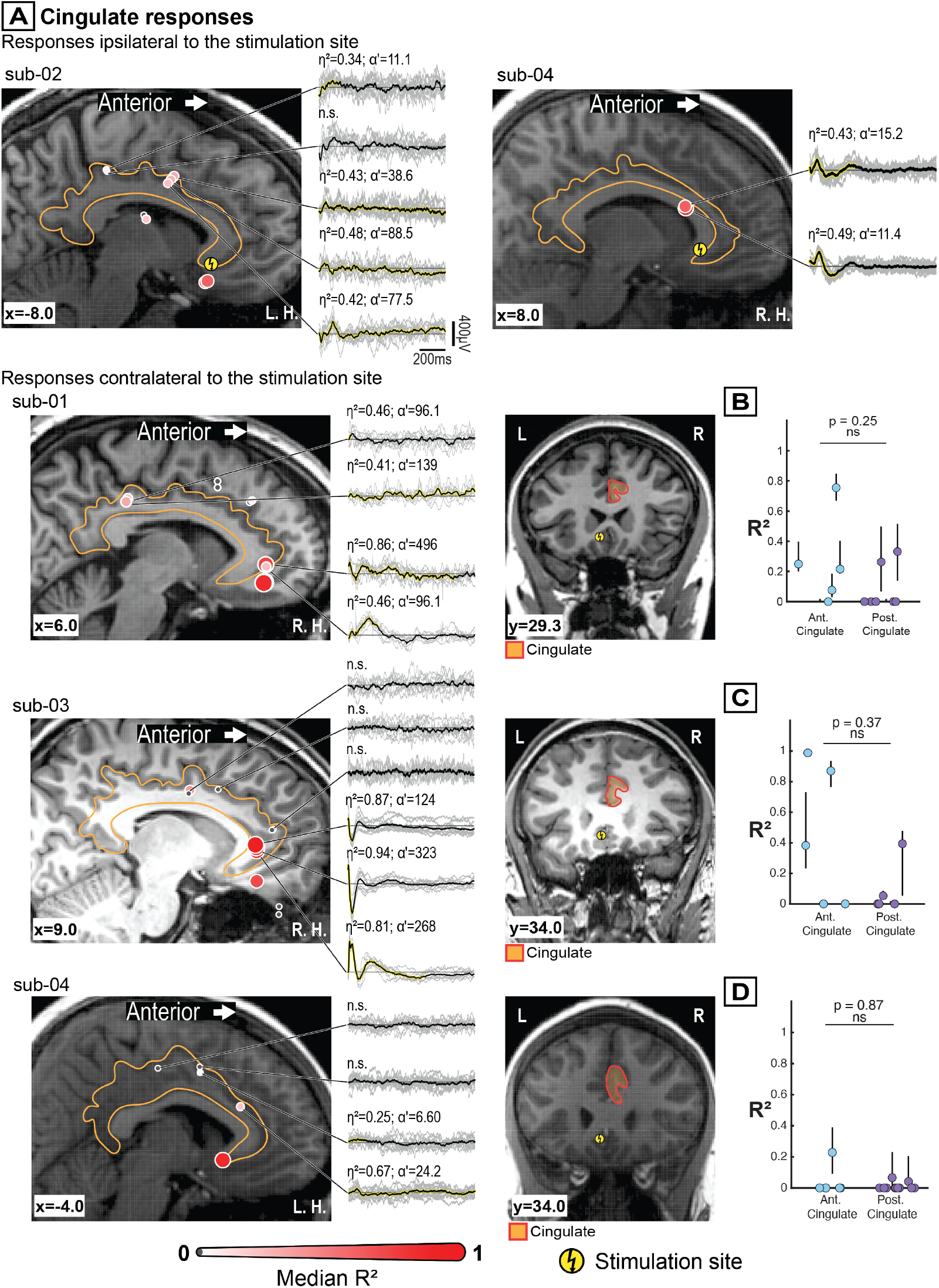
Cingulate responses after subgenual cingulate stimulation. **A**. Brain stimulation evoked potentials (BSEPs) recorded from cingulate electrodes ipsilateral (subjects 2 and 4) and contralateral (subjects 1, 3, and 4) to the subgenual cingulate (sgCC) stimulation site. Significant responses were observed in both hemispheres. **B–D**. Comparison of response strength (median *R*^2^) between anterior and posterior cingulate recording sites upon stimulation of contralateral sgCC for individual subjects. No significant differences were observed between anterior and posterior cingulate contacts (Mann–Whitney U test, subject 1: p = 0.25; subject 3: p = 0.37; subject 4: p = 0.87). Individual points represent median *R*^2^ in the recording channels, with error bars indicating interquartile range across trials.

**Supplementary Figure 4.**
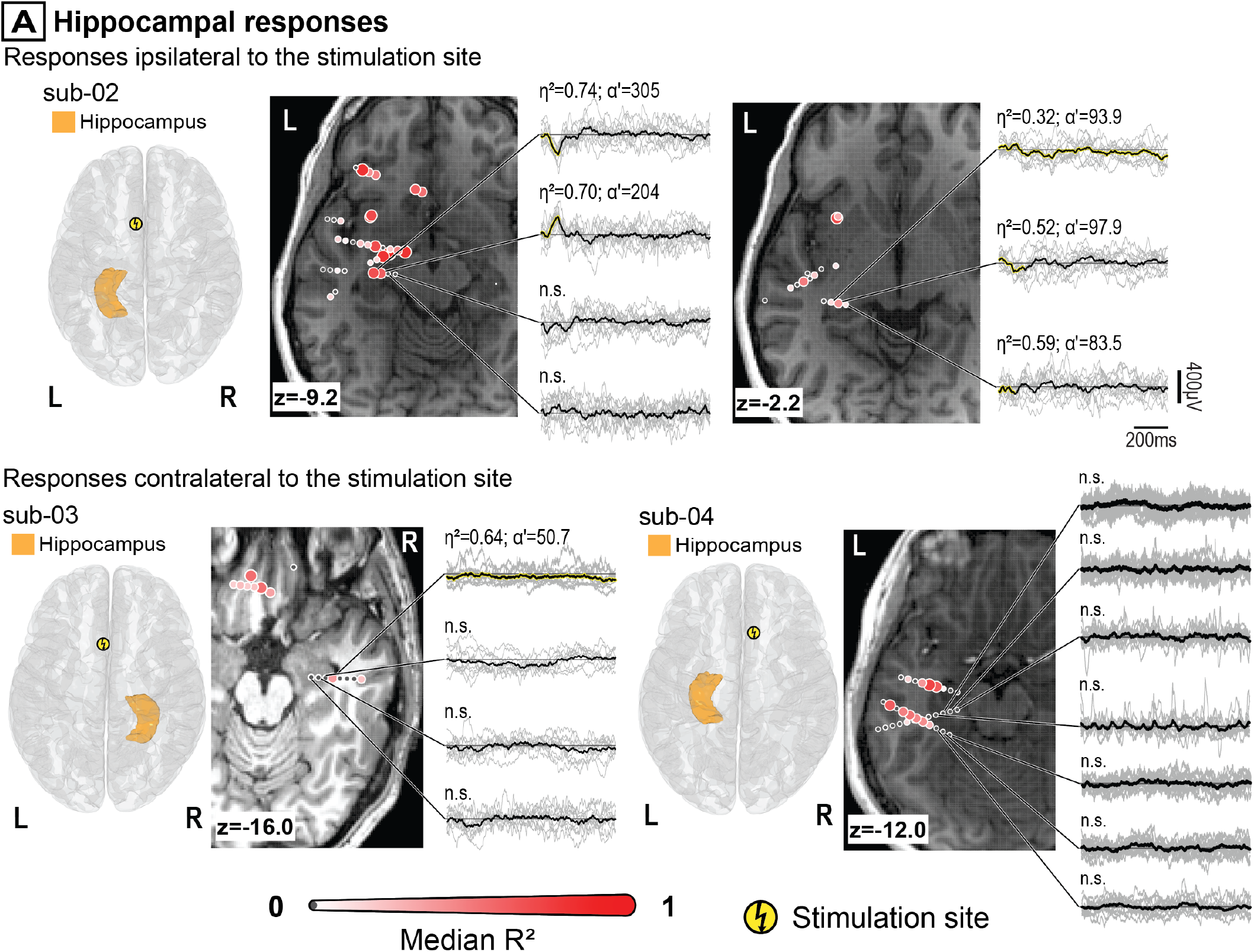
Hippocampal responses after subgenual cingulate stimulation. **A**. Brain stimulation evoked potentials (BSEPs) recorded from hippocampal electrodes ipsilateral (subject 2) and contralateral (subjects 3 and 4) to the subgenual cingulate (sgCC) stimulation site. Multiple ipsilateral hippocampal contacts demonstrated significant responses with relatively high response consistency and larger-amplitude waveforms, whereas contralateral hippocampal contacts showed fewer significant responses and predominantly weak or non-significant activity. These findings support a predominantly ipsilateral propagation of sgCC-evoked activity to the hippocampus.

**Supplementary Figure 5.**
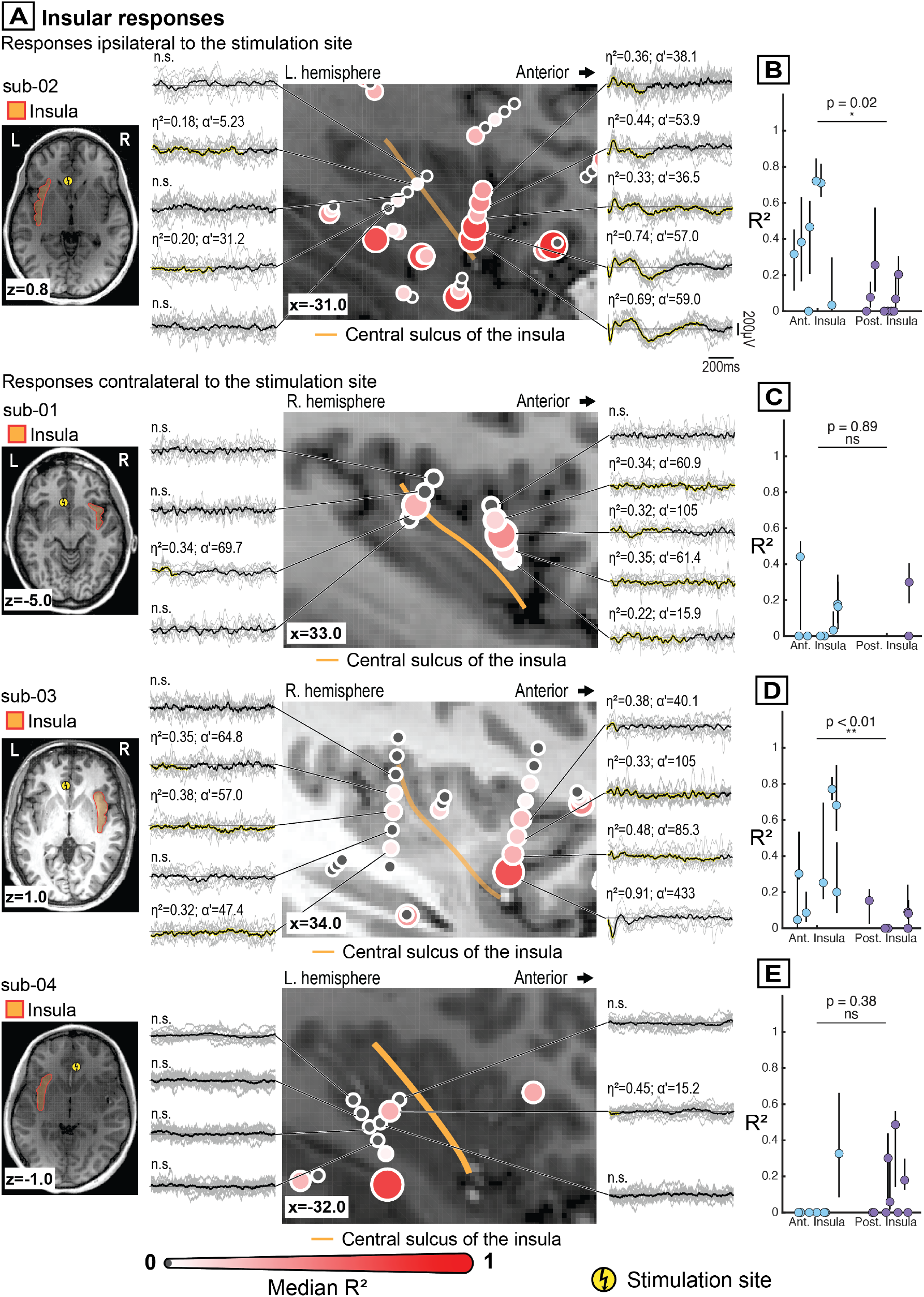
Insular responses after subgenual cingulate stimulation. **A**.Stimulation-evoked responses recorded from insular electrodes ipsilateral (subject 2) and contralateral (subjects 1, 3, and 4) to the sgCC stimulation site are shown together with electrode locations relative to the central insular sulcus, which divides the insula into anterior and posterior portions. In the single subject with ipsilateral insular recordings, sgCC stimulation elicited stronger responses in the anterior than the posterior insula. **B-E**. Comparison of response strength (median *R*^2^) between anterior and posterior insular recording sites for individual subjects. Ipsilateral responses in subject 2 were stronger in anterior than posterior insula (**B**, Mann–Whitney U test, p = 0.02), whereas contralateral responses were variable across subjects, with no significant difference in subject 1 (**C**, Mann– Whitney U test, p = 0.89) or subject 4 (**E**, p = Mann–Whitney U test, 0.38), and stronger anterior responses in subject 3 (**D**, Mann–Whitney U test, p < 0.01). Individual points represent median *R*^2^ values for recording channels, with error bars indicating the interquartile range across trials.

## Supplementary Table

**Supplementary Table 1.** SPES of the sgCC elicited reproducible and spatially organized BSEPs spanning frontal, limbic, and paralimbic networks.

| Subject | Significant channels (%) | Mean $\tau_R \pm \text{std}$ | Median $R^2$ (IQR) | Mean $\eta \pm \text{std}$ | Mean $\alpha' \pm \text{std}$ |
| --- | --- | --- | --- | --- | --- |
| **Orbitofrontal cortex, ipsilateral stimulation** |  |  |  |  |  |
| Sub-01 | 4/5 (80%) | 0.56 ± 0.53 | 0.57 (0.43) | 0.55 ± 0.21 | 108.98 ± 65.39 |
| Sub-03 | 8/10 (80%) | 0.76 ± 0.60 | 0.22 (0.06) | 0.64 ± 0.07 | 52.11 ± 9.87 |
| **Orbitofrontal cortex, contralateral stimulation** |  |  |  |  |  |
| Sub-01 | 2/9 (22.22%) | 0.27 ± 0.28 | 0.42 (0.04) | 0.46 ± 0.11 | 40.97 ± 54.47 |
| Sub-03 | 4/17 (23.53%) | 1.38 ± 0.15 | 0.18 (0.19) | 0.48 ± 0.19 | 45.88 ± 8.65 |
| **Ventromedial prefrontal cortex, ipsilateral stimulation** |  |  |  |  |  |
| Sub-03 | 4/4 (100.00%) | 0.80 ± 0.52 | 0.66 (0.44) | 0.58 ± 0.22 | 93.81 ± 55.17 |
| Sub-02 | 2/2 (100.00%) | 0.81 ± 0.87 | 0.59 (0.06) | 0.61 ± 0.12 | 115.81 ± 36.12 |
| **Ventromedial prefrontal cortex, contralateral stimulation** |  |  |  |  |  |
| Sub-01 | 4/4 (100.00%) | 0.45 ± 0.29 | 0.82 (0.40) | 0.73 ± 0.32 | 313.95 ± 187.49 |
| Sub-03 | 3/4 (75.00%) | 0.44 ± 0.41 | 0.36 (0.42) | 0.57 ± 0.16 | 92.37 ± 38.09 |
| Sub-04 | 2/4 (50.00%) | 0.11 ± 0.00 | 0.92 (0.00) | 0.78 ± 0.06 | 27.44 ± 11.67 |
| **Anterior cingulate cortex, ipsilateral stimulation** |  |  |  |  |  |
| Sub-01 | 3/3 (100.00%) | 0.30 ± 0.07 | 0.69 (0.20) | 0.66 ± 0.05 | 126.23 ± 68.03 |
| Sub-04 | 3/4 (75.00%) | 0.42 ± 0.21 | 0.36 (0.46) | 0.42 ± 0.20 | 12.09 ± 10.84 |
| **Posterior cingulate cortex, ipsilateral stimulation** |  |  |  |  |  |
| Sub-02 | 5/6 (83.33%) | 0.88 ± 0.65 | 0.25 (0.11) | 0.40 ± 0.06 | 55.63 ± 31.13 |
| Sub-04 | 1/6 (16.67%) | 0.16 ± 0.00 | 0.04 (0.00) | 0.67 ± 0.00 | 24.16 ± 0.00 |
| **Anterior cingulate cortex, contralateral stimulation** |  |  |  |  |  |
| Sub-01 | 4/5 (80.00%) | 0.65 ± 0.49 | 0.23 (0.36) | 0.53 ± 0.27 | 182.74 ± 211.70 |
| Sub-03 | 3/5 (60.00%) | 0.32 ± 0.29 | 0.87 (0.45) | 0.73 ± 0.25 | 460.81 ± 287.58 |
| Sub-04 | 1/6 (16.67%) | 1.43 ± 0.00 | 0.23 (0.00) | 0.25 ± 0.00 | 6.60 ± 0.00 |
| **Posterior cingulate cortex, contralateral stimulation** |  |  |  |  |  |
| Sub-01 | 2/7 (28.57%) | 0.69 ± 0.90 | 0.30 (0.07) | 0.44 ± 0.04 | 117.36 ± 30.05 |
| Sub-03 | 2/5 (40.00%) | 0.87 ± 0.70 | 0.22 (0.34) | 0.57 ± 0.12 | 89.97 ± 2.40 |
| Sub-04 | 2/11 (18.18%) | 0.11 ± 0.06 | 0.05 (0.03) | 0.70 ± 0.05 | 29.47 ± 7.51 |
| **Amygdala, ipsilateral stimulation** |  |  |  |  |  |
| Sub-02 | 8/9 (88.89%) | 0.39 ± 0.48 | 0.59 (0.62) | 0.63 ± 0.22 | 47.17 ± 41.04 |
| **Amygdala, contralateral stimulation** |  |  |  |  |  |
| Sub-04 | 1/3 (33.33%) | 0.55 ± 0.00 | 0.06 (0.00) | 0.24 ± 0.00 | 0.10 ± 0.00 |
| **Hippocampus, ipsilateral stimulation** |  |  |  |  |  |
| Sub-02 | 5/7 (71.43%) | 0.38 ± 0.52 | 0.42 (0.51) | 0.57 ± 0.17 | 156.90 ± 96.26 |
| **Hippocampus, contralateral stimulation** |  |  |  |  |  |
| Sub-03 | 1/8 (12.50%) | 1.35 ± 0.00 | 0.37 (0.00) | 0.64 ± 0.00 | 50.67 ± 0.00 |
| Sub-04 | 0/6 (0.00%) | NaN ± NaN | NaN (NaN) | NaN ± NaN | NaN ± NaN |
| **Anterior insular cortex, ipsilateral stimulation** |  |  |  |  |  |
| Sub-02 | 6/7 (85.71%) | 0.70 ± 0.43 | 0.42 (0.39) | 0.46 ± 0.22 | 41.62 ± 20.26 |
| **Posterior insular cortex, ipsilateral stimulation** |  |  |  |  |  |
| Sub-02 | 4/9 (44.44%) | 0.73 ± 0.56 | 0.14 (0.16) | 0.34 ± 0.14 | 45.49 ± 18.74 |
| **Anterior insular cortex, contralateral stimulation** |  |  |  |  |  |
| Sub-01 | 4/8 (50.00%) | 0.96 ± 0.46 | 0.17 (0.21) | 0.31 ± 0.06 | 60.72 ± 36.26 |
| Sub-03 | 7/7 (100.00%) | 0.40 ± 0.36 | 0.25 (0.47) | 0.56 ± 0.19 | 135.54 ± 138.07 |
| Sub-04 | 1/7 (14.29%) | 0.54 ± 0.0 | 0.33 (0.00) | 0.29 ± 0.00 | 11.19 ± 0.00 |
| **Posterior insular cortex, contralateral stimulation** |  |  |  |  |  |
| Sub-01 | 1/2 (50.00%) | 0.23 ± 0.00 | 0.30 (0.00) | 0.34 ± 0.00 | 69.75 ± 0.00 |
| Sub-03 | 3/7 (42.86%) | 0.99 ± 0.58 | 0.09 (0.05) | 0.35 ± 0.03 | 56.44 ± 8.71 |
| Sub-04 | 4/9 (44.44%) | 0.43 ± 0.59 | 0.24 (0.27) | 0.43 ± 0.16 | 9.67 ± 3.79 |

| Temporal pole, ipsilateral stimulation |  |  |  |  |  |
| --- | --- | --- | --- | --- | --- |
| Sub-02 | 4/9 (44.44%) | 0.31 ± 0.10 | 0.31 (0.36) | 0.54 ± 0.07 | 54.31 ± 27.38 |
| Temporal pole, contralateral stimulation |  |  |  |  |  |
| Sub-04 | 3/5 (60.00%) | 0.67 ± 0.60 | 0.13 (0.30) | 0.35 ± 0.10 | 76.70 ± 54.28 |

